# Proportion and Determinants of Cervical Cancer Knowledge Among Female Sex Workers in Kilimanjaro: A Community-Based Cross-Sectional Study guided by the Socioecological Model

**DOI:** 10.1101/2025.05.01.25326803

**Authors:** Gumbo D. Silas, Alex Mremi, Innocent H. Uggh, Bariki Mchome, Alma R. Nzunda, Gaudensia Olomi, Prisca Marandu, Leah Mmari, Happiness Kilamwai, Blandina T. Mmbaga, Patricia Swai

**Affiliations:** KCMC University, Moshi, Tanzania; Kilimanjaro Christian Medical Center, Moshi Tanzania; Kilimanjaro Clinical Research Institute, Moshi, Tanzania; Global & Planetary Health Working Group, Martin-Luther-University Halle-Wittenberg, Germany

**Keywords:** Cervical Cancer, Female Sex Workers, Cervical cancer Knowledge, Prevention

## Abstract

**Background:** Cervical cancer remains a significant public health issue in Tanzania, with high morbidity and mortality rates largely driven by limited awareness and access to preventive services. Female sex workers (FSWs) are particularly vulnerable due to their increased exposure to HPV through multiple sexual partners. This study aimed to assess the level of cervical cancer knowledge and associated factors among FSWs in Kilimanjaro region.

**Methods:** A community-based cross-sectional study was conducted among 351 FSWs aged 25-49 years in hotspot areas of Kilimanjaro region between June and July 2024. Respondent-driven sampling (RDS) was employed. Data were collected using an interviewer-administered questionnaire. Statistical analysis was performed with SPSS version 27.0, and Poisson regression with robust error variance was used to assess associations. Variables with a p-value less than 0.05 in the multivariate analysis were considered statistically significant.

**Results:** The mean age of study participants was 36.11±5.24 years, with 49.3% aged 25–34 years. The majority (66.1%) resided in urban areas. About 52.1% had attained primary education and only 2.6% had a college or university education. Additionally, 69.5% had at least two children, and 49.9% were separated or divorced.

Among the 351 FSWs studied, 89.0% were aware of cervical cancer. However, only 19.9% had a high level of knowledge about the disease and its prevention, with only 31.2%, 27.8% and 34.5% demonstrating high knowledge of risk factors, symptoms and signs, and preventive measures, respectively. Key factors significantly associated with cervical cancer knowledge included residence (APR:1.17, 95% CI:1.12-1.35), age at sexual debut (APR: 1.15, 95% CI: 1.05-1.26), and (APR: 1.18, 95% CI: 1.09-1.27), age at starting sex work (APR: 1.38, 95% CI: 1.12-1.69) and alcohol consumption (APR: 1.08, 95% CI: 1.002-1.16).

**Conclusion:** Despite high awareness of cervical cancer among FSWs, comprehensive knowledge about its risk factors, symptoms, and prevention remains low. These findings highlight the urgent need for targeted educational interventions that consider differences in residence, age at sexual debut, age at starting commercial sex, and alcohol consumption.

## BACKGROUND

Cervical cancer remains a significant global public health concern, with an estimated 662,000 new cases and 349,000 deaths reported in 2022[1]. The burden is disproportionately high in Sub-Saharan Africa, where over 94% of cases and deaths occur [1]. Tanzania is among the most affected countries in the region, recording 10,868 cases and 6,832 deaths in 2022 [2]. The high burden of cervical cancer is primarily driven by persistent infection with high-risk carcinogenic human papillomavirus (HPV), compounded by poor awareness, misconceptions, and limited access to preventive services, leading to low screening uptake [3],[4] Awareness of cervical cancer symptoms, risk factors, and preventive measures plays a crucial role in early health-seeking behaviour. Women with higher knowledge are more likely to undergo regular screening, which facilitates early detection and treatment, ultimately reducing mortality rates [5][6].

Several studies have consistently highlighted low knowledge levels about cervical cancer in the general population of women. For instance, a study conducted in China found inadequate awareness among women[7]. In India, a study involving 7,688 women reported that only 40.22% had overall knowledge of cervical cancer, while awareness of risk factors, symptoms, and signs was even lower (23.01%)[8]. Similarly, in Ethiopia, a study of 520 women found that only 27.7% had adequate knowledge[9].

In Tanzania, despite the availability of HPV vaccination and cervical cancer screening, many women remain at risk due to limited access to screening services and insufficient knowledge [10]. A study in Northern Tanzania among 2,192 women found that only 22% had a high level of knowledge about cervical cancer[10]. Misconceptions, fear, stigma, and cultural barriers further hinder screening uptake, emphasizing the need for comprehensive awareness programs[11].

Among high-risk populations, female sex workers (FSWs) are particularly vulnerable to cervical cancer due to increased exposure to HPV infection, high-risk sexual behaviors, and barriers to accessing healthcare services. Studies indicate that FSWs have higher HPV infection rates than women in the general population, making targeted prevention strategies essential. However, little is known about their level of awareness, knowledge of cervical cancer, and screening behaviors in Tanzania. Understanding the extent of cervical cancer knowledge and its associated factors among this high-risk group is crucial for designing targeted interventions that enhance awareness, increase screening uptake, and promote early detection. This study aims to determine the level of cervical cancer knowledge and associated factors among FSWs in Kilimanjaro region.

The findings will provide evidence-based recommendations for designing effective health education programs, improving outreach strategies, and enhancing cervical cancer prevention efforts tailored to FSWs. Moreover, the study aligns with national and global efforts to reduce cervical cancer morbidity and mortality. It also supports the United Nations Sustainable Development Goals (SDGs), particularly SDG 3, which promotes good health and well-being, and SDG 5, which aims to achieve gender equality and empower women by improving access to life-saving preventive services.

### Theoretical Framework

This study is guided by the Social Ecological Model (SEM), which explains the multi-level determinants of knowledge about cervical cancer. This framework highlights that knowledge of cervical cancer is shaped by both direct and indirect influences across multiple levels. Understanding these interconnections can help design targeted interventions to improve cervical cancer awareness and screening uptake among FSWs.[12]

The model consists of five levels:

1. **Individual Level**: Includes factors such as education level, age at sexual debut, age at starting sex work, alcohol consumption, and personal health experiences. These factors directly shape a person’s knowledge of cervical cancer.[13]
2. **Interpersonal Level**: Includes the influence of healthcare providers and peer networks. Positive interactions with healthcare workers and peer discussions can enhance knowledge and awareness.
3. **Institutional/Organizational Level**: Workplace policies and health education programs. The presence of cervical cancer screening services, training for healthcare providers, and institutional policies on reproductive health affect knowledge dissemination.[14]
4. **Community Level**: Includes residence, availability of health services, and media exposure. These factors influence access to health education and screening information.[15]
5. **Societal Level**: Includes stigma, cultural beliefs, healthcare policies, and access to screening programs. These factors indirectly shape knowledge by influencing healthcare accessibility and the dissemination of information.[16]

## METHODOLOGY

### Study design, study population, and eligibility criteria

This was a community-based cross-sectional study which was conducted among female sex workers aged 25-49 years in the hotspots located in Moshi district council and Moshi municipality in Kilimanjaro region.

### Study area

Kilimanjaro region is among 26 regions of Tanzania’s mainland. It is in northern Tanzania. It borders Kenya to the north, Tanga region to the northeast, Arusha to the west, and Manyara region to the south. The dominant ethnic group is Chaga and pare, and the major economic activities are tourism, agriculture, and business. It is the home for Mount Kilimanjaro, the highest mountain in Africa.

### Sample size and sampling techniques

Participants of this study were enrolled through the respondent-driven sampling technique (RDS). RDS is a network-based sampling technique used to study hard-to-reach or hidden populations, where traditional random sampling methods may not be effective or feasible. [17]. With RDS the recruitment of study participants is usually a wave based. It begins with recruitment of a few participants called seeds. In this study, the seeds were the well-trained peer educators of FSWs. In first wave four seeds were provided with four coupons each to distribute to their peers. Each one of the newly recruited participants (Wave 1) were also provided the four coupons to distribute to their peers (wave 2), then the process continued until saturation of the desired sample size was reached.

The sample size of 351 was estimated using the formula below, where **Z is** the z-score for 95% confidence level, **P** is the proportion of FHWs with high knowledge of cervical cancer, **1-P is** the proportion of FSWs who had low knowledge, and **ꜫ is** the margin of error or precision level.

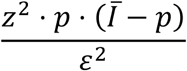

### Pilot study and study variables

To ensure this study’s quality, reliability, and feasibility, a pilot study was conducted among 10% of the study participants who were not included in this study. The dependent variable was the level of knowledge of cervical cancer, while the independent variables included the sociodemographic and clinical characteristics of study participants.

### Data collection tool and procedure

An interviewer-administered questionnaire was used to collect data on the sociodemographic and clinical characteristics of study participants. Trained research assistants were supported by peer educators in reaching the hotspots. Participants were interviewed after consenting to participate in the study.

### Data management and analysis

Data were cross-checked for accuracy, quality, and errors. The analysis was performed using SPSS version 27.0, where in descriptive statistics, categorical variables were summarized in frequencies and percentages while continuous variables were summarised into mean and standard deviation. In inferential statistics, the Chi-square test was performed to assess the association between the independent variables and level of knowledge of cervical cancer, and Poisson Regression with Robust Error Variance was performed to determine the magnitude of the observed associations at a 95% statistical significance level with a p-value cut-off point of 0.05. Regarding the analysis of knowledge levels, the standardized mean score for knowledge of risk factors was 0.4475, and the total standardized mean score was 0.337, with knowledge levels classified as follows: Scores ≤ 0.337: Low knowledge; and Scores > 0.337: High knowledge.

## RESULTS

### Sociodemographic characteristics of study participants

The study involved 351 female sex workers in Kilimanjaro region, with a mean age of 36.11±5.24 years. The majority (49.3%) were aged 25–34 years, while 7.1% were in the 45– 49-year age group. Most participants resided in urban areas (66.1%), while 33.9% were from rural settings. More than half (52.1%) had attained primary education, while 41.6% had completed secondary education. A small proportion (3.7%) had never attended school, and 2.6% had a college or university education. Nearly half (49.9%) were separated or divorced, while 31.6% were single and 18.5% were married or cohabiting. Most participants (69.5%) had given birth to at least two children (Table 1).

**Table 1:**
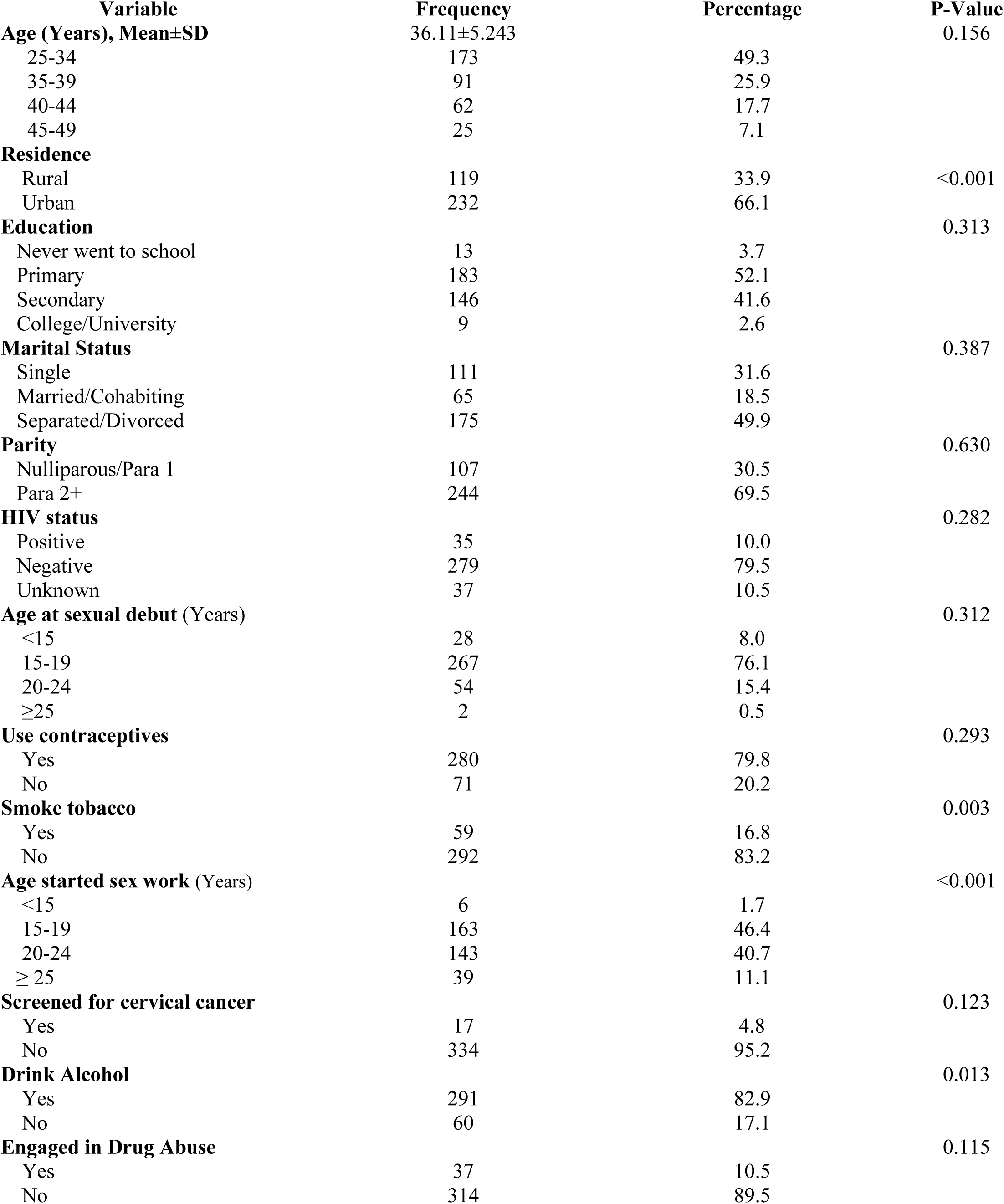
Sociodemographic, sexual and behavioural characteristics of FSWs in Kilimanjaro region (N=351)

| Variable | Frequency | Percentage | P-Value |
| --- | --- | --- | --- |
| **Age (Years), Mean±SD** | 36.11±5.243 |  | 0.156 |
| 25-34 | 173 | 49.3 |  |
| 35-39 | 91 | 25.9 |  |
| 40-44 | 62 | 17.7 |  |
| 45-49 | 25 | 7.1 |  |
| **Residence** |  |  |  |
| Rural | 119 | 33.9 | <0.001 |
| Urban | 232 | 66.1 |  |
| **Education** |  |  | 0.313 |
| Never went to school | 13 | 3.7 |  |
| Primary | 183 | 52.1 |  |
| Secondary | 146 | 41.6 |  |
| College/University | 9 | 2.6 |  |
| **Marital Status** |  |  | 0.387 |
| Single | 111 | 31.6 |  |
| Married/Cohabiting | 65 | 18.5 |  |
| Separated/Divorced | 175 | 49.9 |  |
| **Parity** |  |  | 0.630 |
| Nulliparous/Para 1 | 107 | 30.5 |  |
| Para 2+ | 244 | 69.5 |  |
| **HIV status** |  |  | 0.282 |
| Positive | 35 | 10.0 |  |
| Negative | 279 | 79.5 |  |
| Unknown | 37 | 10.5 |  |
| **Age at sexual debut (Years)** |  |  | 0.312 |
| <15 | 28 | 8.0 |  |
| 15-19 | 267 | 76.1 |  |
| 20-24 | 54 | 15.4 |  |
| ≥25 | 2 | 0.5 |  |
| **Use contraceptives** |  |  | 0.293 |
| Yes | 280 | 79.8 |  |
| No | 71 | 20.2 |  |
| **Smoke tobacco** |  |  | 0.003 |
| Yes | 59 | 16.8 |  |
| No | 292 | 83.2 |  |
| **Age started sex work (Years)** |  |  | <0.001 |
| <15 | 6 | 1.7 |  |
| 15-19 | 163 | 46.4 |  |
| 20-24 | 143 | 40.7 |  |
| ≥ 25 | 39 | 11.1 |  |
| **Screened for cervical cancer** |  |  | 0.123 |
| Yes | 17 | 4.8 |  |
| No | 334 | 95.2 |  |
| **Drink Alcohol** |  |  | 0.013 |
| Yes | 291 | 82.9 |  |
| No | 60 | 17.1 |  |
| **Engaged in Drug Abuse** |  |  | 0.115 |
| Yes | 37 | 10.5 |  |
| No | 314 | 89.5 |  |

About 76.1% had their first sexual experience between 15–19 years, while 8.0% had an early sexual debut before 15 years. The age of initiating sex work varied, with 46.4% starting between 15–19 years, and 40.7% between 20–24 years. In total, 10.0% were HIV-positive, while 79.5% tested negative, and 10.5% were unaware of their status. The majority (79.8%) reported using contraceptives. Substance use was notable, with 82.9% consuming alcohol, 16.8% smoking tobacco, and 10.5% engaging in drug abuse. (Table 1)

### FSWs’ Knowledge test scores on Causes and Risk Factors of cervical cancer

The study assessed FSWs’ knowledge scores on cervical cancer with a Cronbach’s Alpha of 0.807, indicating good internal consistency. When asked whether HPV infection causes cervical cancer, only 7.7% (27) strongly agreed, while 42.7% (150) were neutral, and 30.5% (107) disagreed. Similarly, smoking cigarettes as a risk factor had a high level of uncertainty, with 47.3% (166) neutral, while only 3.4% (12) strongly agreed. (Table 2) Regarding HIV infection, 48.4% (170) were neutral, while 25.4% (89) disagreed, and only 2.8% (10) strongly agreed. The use of contraceptive pills for a long time was also met with uncertainty, as 38.7% (136) were neutral, 29.9% (105) disagreed, and 6.0% (21) strongly agreed. Other risk factors, such as having other STIs (40.2%, 141 neutral), an uncircumcised partner (41.0%, 144 neutral), early sexual debut (41.0%, 144 neutral), and multiple sexual partners (42.2%, 148 neutral), showed similar patterns of limited agreement. Not going for regular screening was acknowledged as a risk factor by 6.6% (23) strongly agreeing, while 44.2% (155) were neutral. Since the standardized mean score for knowledge of risk factors was 0.4475, this indicates a relatively better understanding of risk factors compared to symptoms. (Table 2)

**Table 2:**
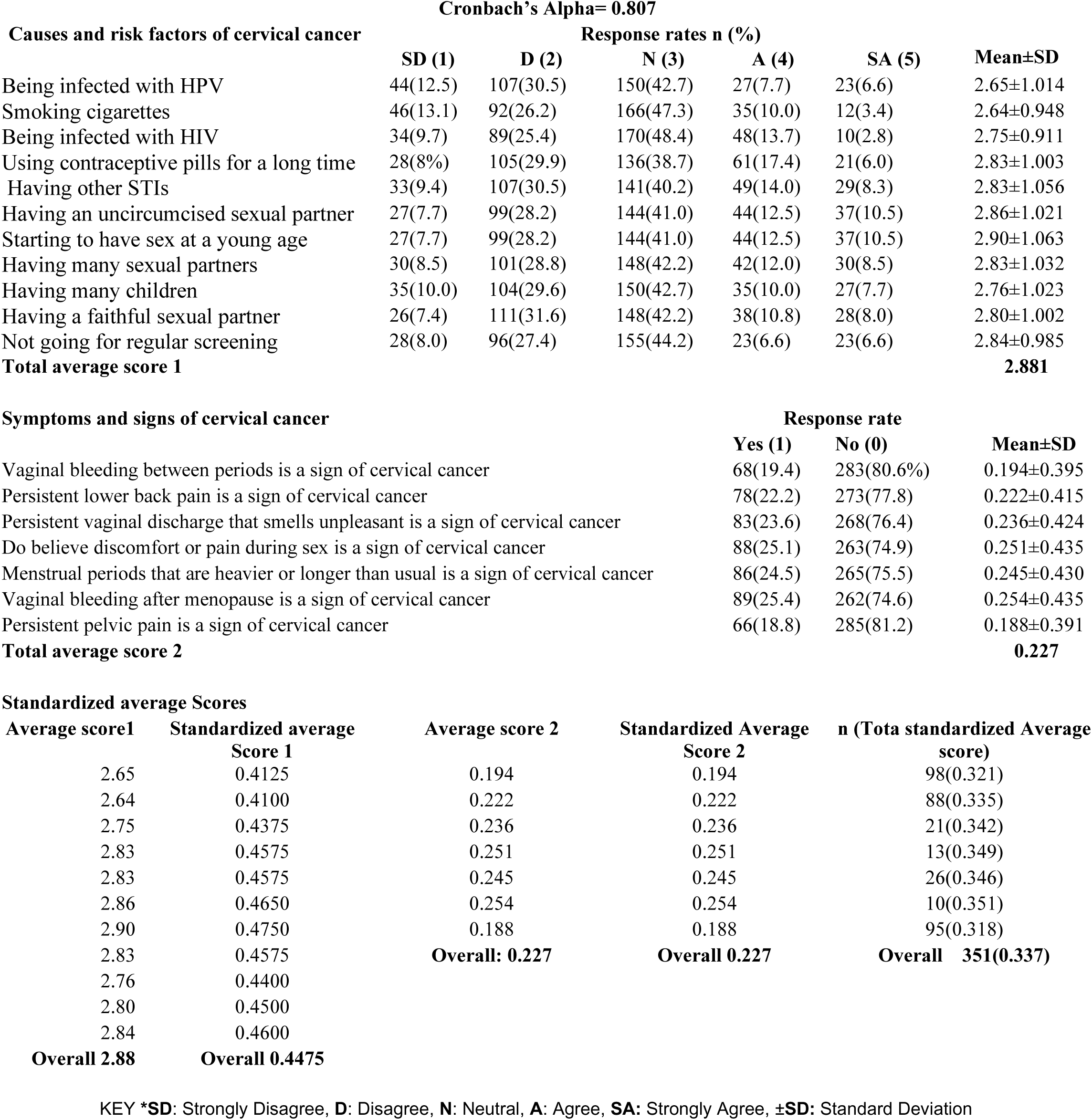
Response rates and standardized scores about knowledge of cervical cancer among FSWs in Kilimanjaro region (N=351)

### FSWs’ Knowledge test scores on Symptoms and Signs of cervical cancer

Knowledge scores about symptoms of cervical cancer were notably low. Only 19.4% (68) correctly identified vaginal bleeding between periods as a symptom, while 80.6% (283) were unaware. Persistent lower back pain was recognized by 22.2% (78), unpleasant vaginal discharge by 23.6% (83), pain during sex by 25.1% (88), heavier/longer menstrual periods by 24.5% (86), and vaginal bleeding after menopause by 25.4% (89). The least recognized symptom was persistent pelvic pain, identified by 18.8% (66), with 81.2% (285) not recognizing it as a symptom. Since the standardized mean score for knowledge of symptoms was 0.227, this reflects poor awareness of cervical cancer symptoms (Table 2).

### Proportion of Knowledge on Cervical Cancer and Its Prevention Among FSWs

Generally, vast majority of participants, 281 (80.1%), had scores less than or equal to 0.337, indicating a low level of knowledge, particularly regarding symptoms. In contrast, only 70 (19.9%) participants scored above 0.337, suggesting a high level of knowledge. (Figure 1)

**Figure 1:**
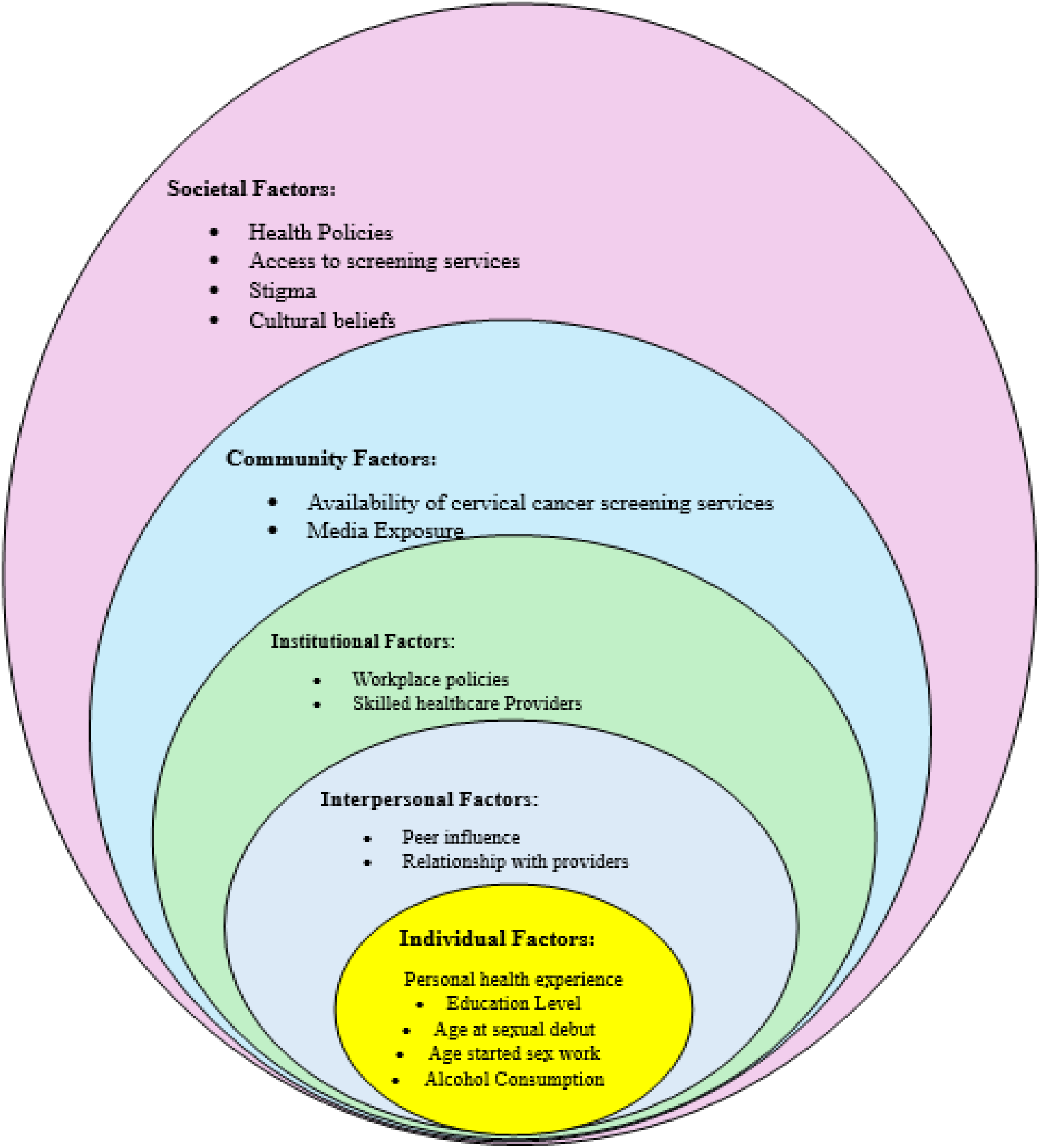
SEM showing factors influencing the level of knowledge about cervical cancer among FSWs.

Age showed a slight increase in knowledge with advancing years, with the highest proportion of high knowledge among those aged 40-44 years (25.8%). Rural residents (36.1%) had significantly higher knowledge levels than urban dwellers (11.6%). Education played a key role, as knowledge increased with higher educational attainment, peaking at 33.3% among those with college/university education. Marital status and parity showed minor variations, with separated/divorced FSWs (21.7%) and those with fewer children (21.5%) having slightly higher knowledge. Additionally, those who were HIV-negative (21.5%) had greater knowledge than their HIV-positive (17.1%) and unknown-status (10.8%) counterparts. (Table 3, Figure 2)

**Figure 2:**
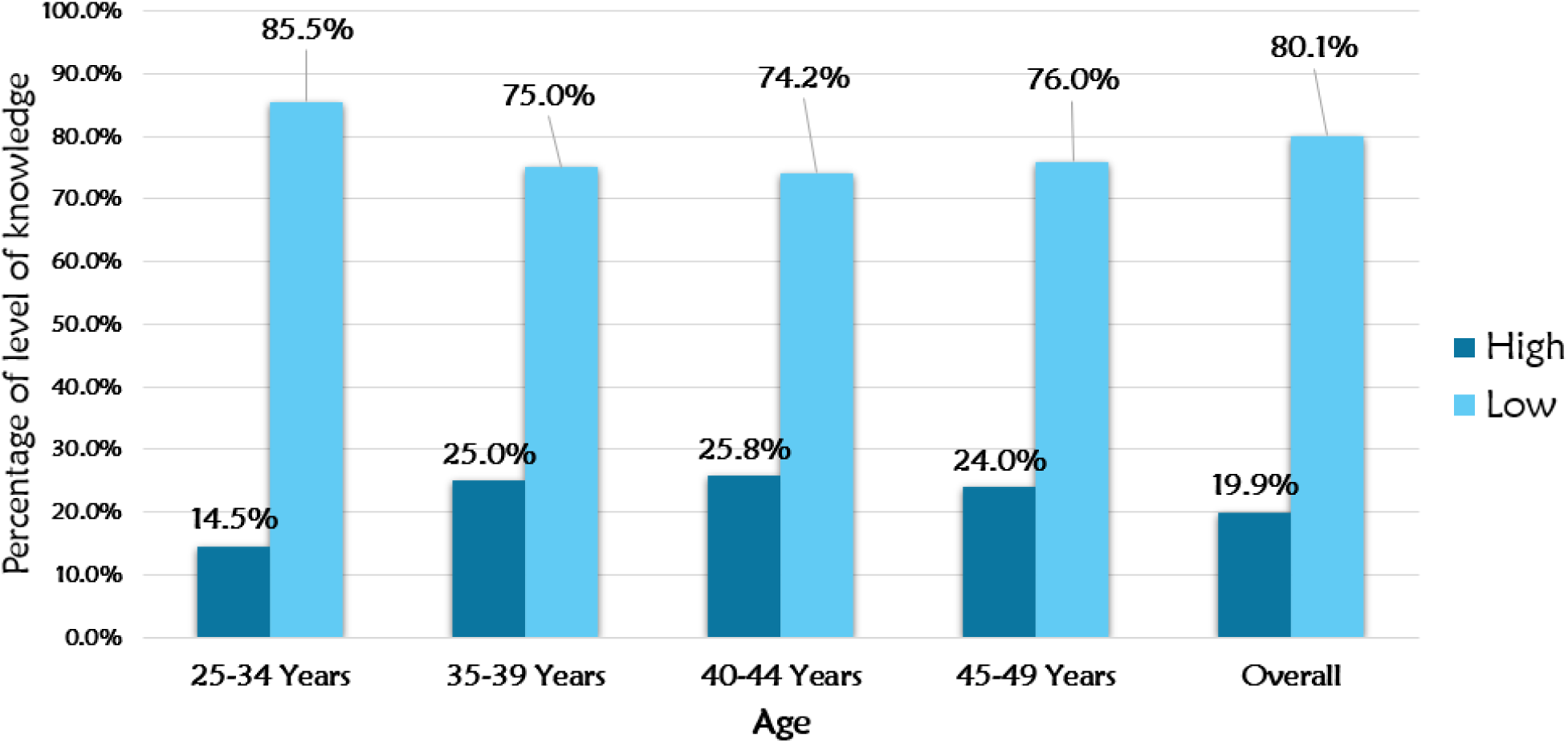
Level of knowledge about cervical cancer among FSWs in Kilimanjaro Region (N=351)

**Table 3:**
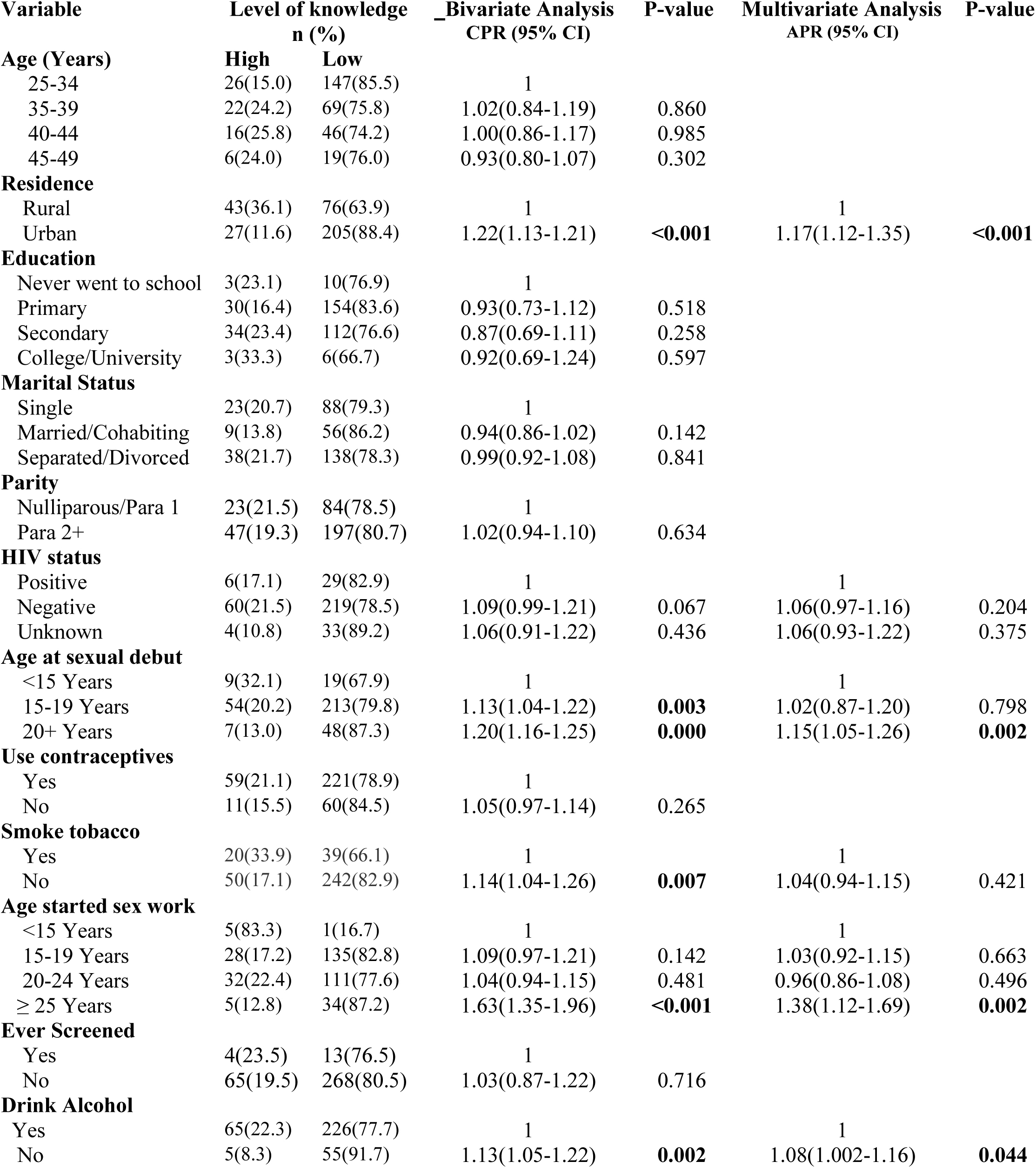
Level of Knowledge of cervical cancer and associated factors among FSWs in Kilimanjaro (N=351)

| Variable | Level of knowledge<br>n (%) |  | _Bivariate Analysis<br>CPR (95% CI) | P-value | Multivariate Analysis<br>APR (95% CI) | P-value |
| --- | --- | --- | --- | --- | --- | --- |
|  | High | Low |  |  |  |  |
| **Age (Years)** |  |  |  |  |  |  |
| 25-34 | 26(15.0) | 147(85.5) | 1 |  |  |  |
| 35-39 | 22(24.2) | 69(75.8) | 1.02(0.84-1.19) | 0.860 |  |  |
| 40-44 | 16(25.8) | 46(74.2) | 1.00(0.86-1.17) | 0.985 |  |  |
| 45-49 | 6(24.0) | 19(76.0) | 0.93(0.80-1.07) | 0.302 |  |  |
| **Residence** |  |  |  |  |  |  |
| Rural | 43(36.1) | 76(63.9) | 1 |  | 1 |  |
| Urban | 27(11.6) | 205(88.4) | 1.22(1.13-1.21) | <0.001 | 1.17(1.12-1.35) | <0.001 |
| **Education** |  |  |  |  |  |  |
| Never went to school | 3(23.1) | 10(76.9) | 1 |  |  |  |
| Primary | 30(16.4) | 154(83.6) | 0.93(0.73-1.12) | 0.518 |  |  |
| Secondary | 34(23.4) | 112(76.6) | 0.87(0.69-1.11) | 0.258 |  |  |
| College/University | 3(33.3) | 6(66.7) | 0.92(0.69-1.24) | 0.597 |  |  |
| **Marital Status** |  |  |  |  |  |  |
| Single | 23(20.7) | 88(79.3) | 1 |  |  |  |
| Married/Cohabiting | 9(13.8) | 56(86.2) | 0.94(0.86-1.02) | 0.142 |  |  |
| Separated/Divorced | 38(21.7) | 138(78.3) | 0.99(0.92-1.08) | 0.841 |  |  |
| **Parity** |  |  |  |  |  |  |
| Nulliparous/Para 1 | 23(21.5) | 84(78.5) | 1 |  |  |  |
| Para 2+ | 47(19.3) | 197(80.7) | 1.02(0.94-1.10) | 0.634 |  |  |
| **HIV status** |  |  |  |  |  |  |
| Positive | 6(17.1) | 29(82.9) | 1 |  | 1 |  |
| Negative | 60(21.5) | 219(78.5) | 1.09(0.99-1.21) | 0.067 | 1.06(0.97-1.16) | 0.204 |
| Unknown | 4(10.8) | 33(89.2) | 1.06(0.91-1.22) | 0.436 | 1.06(0.93-1.22) | 0.375 |
| **Age at sexual debut** |  |  |  |  |  |  |
| <15 Years | 9(32.1) | 19(67.9) | 1 |  | 1 |  |
| 15-19 Years | 54(20.2) | 213(79.8) | 1.13(1.04-1.22) | 0.003 | 1.02(0.87-1.20) | 0.798 |
| 20+ Years | 7(13.0) | 48(87.3) | 1.20(1.16-1.25) | 0.000 | 1.15(1.05-1.26) | 0.002 |
| **Use contraceptives** |  |  |  |  |  |  |
| Yes | 59(21.1) | 221(78.9) | 1 |  |  |  |
| No | 11(15.5) | 60(84.5) | 1.05(0.97-1.14) | 0.265 |  |  |
| **Smoke tobacco** |  |  |  |  |  |  |
| Yes | 20(33.9) | 39(66.1) | 1 |  | 1 |  |
| No | 50(17.1) | 242(82.9) | 1.14(1.04-1.26) | 0.007 | 1.04(0.94-1.15) | 0.421 |
| **Age started sex work** |  |  |  |  |  |  |
| <15 Years | 5(83.3) | 1(16.7) | 1 |  | 1 |  |
| 15-19 Years | 28(17.2) | 135(82.8) | 1.09(0.97-1.21) | 0.142 | 1.03(0.92-1.15) | 0.663 |
| 20-24 Years | 32(22.4) | 111(77.6) | 1.04(0.94-1.15) | 0.481 | 0.96(0.86-1.08) | 0.496 |
| ≥ 25 Years | 5(12.8) | 34(87.2) | 1.63(1.35-1.96) | <0.001 | 1.38(1.12-1.69) | 0.002 |
| **Ever Screened** |  |  |  |  |  |  |
| Yes | 4(23.5) | 13(76.5) | 1 |  |  |  |
| No | 65(19.5) | 268(80.5) | 1.03(0.87-1.22) | 0.716 |  |  |
| **Drink Alcohol** |  |  |  |  |  |  |
| Yes | 65(22.3) | 226(77.7) | 1 |  | 1 |  |
| No | 5(8.3) | 55(91.7) | 1.13(1.05-1.22) | 0.002 | 1.08(1.002-1.16) | 0.044 |

Behavioural factors also influence knowledge levels. FSWs who started sex work before 15 years had the highest knowledge (83.3%), while those who initiated sex work later had lower knowledge. Tobacco smokers (33.9%) had significantly higher knowledge compared to non-smokers (17.1%), and contraceptive users (21.1%) were slightly more knowledgeable than non-users (15.5%). Alcohol consumption was associated with higher knowledge levels (22.3%) compared to non-drinkers (8.3%). Screening history had minimal impact, with 23.5% of those ever screened having high knowledge compared to 19.5% of those never screened. These findings highlight the need for targeted educational interventions, especially among urban FSWs, those with lower education, and those who started sex work at an older age(Table 3).

### Factors Associated with Cervical Cancer Knowledge Among Female Sex Workers

In this study, residence, age at sexual debut, age at starting sex work, and alcohol consumption were significantly associated with cervical cancer knowledge among this population. Urban dwellers had a lower likelihood of high cervical cancer knowledge compared to rural dwellers (APR: 1.17, 95% CI: 1.12-1.35). Age at sexual debut was also significant, with those who initiated sex at 20-24 years (APR: 1.15, 95% CI: 1.05-1.26,) and ≥25 years (APR: 1.18, 95% CI: 1.09-1.27) being more knowledgeable than those who started before 15 years. Additionally, age at starting sex work played a role, as those who started at 25 years or older had higher knowledge compared to those who began before 15 years (APR: 1.38, 95% CI: 1.12-1.69). Lastly, alcohol consumption was associated with higher cervical cancer knowledge (APR: 1.08, 95% CI: 1.002-1.16) (Table 3).

## DISCUSSION

This study revealed that the majority (88.9%) of female sex workers (FSWs) were aware of cervical cancer and its prevention. However, despite this high awareness, only 19.9% demonstrated a high level of knowledge about cervical cancer and its prevention. These findings highlight a critical gap between awareness and in-depth understanding of the disease, which may have implications for preventive behaviors.

One possible explanation for the low proportion of FSWs with high knowledge is the asymptomatic nature of cervical cancer in its early stages, which may contribute to a low perceived risk of the disease. Without visible symptoms, many individuals may not recognize the urgency of seeking information or preventive measures, such as screening. Additionally, the lack of targeted educational campaigns tailored to this high-risk population could further limit their knowledge and engagement with cervical cancer prevention strategies.

Our findings on awareness are consistent with studies conducted in other regions. For instance, in Thailand, 85.3% of FSWs reported being aware of cervical cancer and its prevention[18]. Similarly, in Abuja, Nigeria, awareness was reported at 71.4%[19]. However, in contrast, significantly lower awareness levels were observed among FSWs in Sokoto State, Nigeria, where only 12.1% of participants were aware of cervical cancer, and just 6.7% knew about its prevention[20]. These discrepancies may be attributed to differences in health education initiatives, access to healthcare services, and sociocultural factors influencing information dissemination.

Regarding knowledge, our findings align with studies showing that many FSWs have limited knowledge about cervical cancer despite high awareness levels. For example, in Shashemene Town, Ethiopia, 50.1% of FSWs had high knowledge of cervical cancer and its prevention[21], In Pakistan, 40% of FSWs were reported to have sufficient knowledge[22]. Similarly, a study in Douala Municipality, Cameroon, found that 35.5% of FSWs had adequate knowledge[23]. Additionally, a study conducted in South Africa found that only 24.3% of FSWs had high knowledge of cervical cancer and its prevention[24]. The lower proportion of FSWs with high knowledge in our study suggests that more efforts are needed to enhance comprehensive education on cervical cancer, including risk factors, symptoms, screening, and vaccination.

These findings emphasize the need for targeted interventions that go beyond raising awareness to improving knowledge and facilitating informed decision-making about cervical cancer prevention. Culturally appropriate and accessible educational campaigns, tailored specifically to FSWs, could help bridge this knowledge gap. Moreover, integrating cervical cancer education into existing sexual and reproductive health services for FSWs may enhance their understanding and uptake of preventive measures such as HPV vaccination and regular screening.

This study also identified several factors associated with cervical cancer knowledge among FSWs, including residence, age at sexual debut, age at which they started engaging in commercial sex, and alcohol consumption. These findings provide valuable insights into the determinants of knowledge levels within this population and highlight areas for targeted interventions.

Residence emerged as a significant factor, with FSWs residing in urban areas having a higher likelihood of possessing greater cervical cancer knowledge (AOR: 1.17). This aligns with findings from elsewhere, [25], which reported that urban residents were more informed about cervical cancer (AOR: 1.365). Urban settings generally offer better access to health information, structured healthcare programs, and media-driven awareness campaigns. In contrast, FSWs in rural areas may face limited exposure to cervical cancer education due to fewer health outreach initiatives and inadequate healthcare infrastructure. These findings emphasise the need to strengthen cervical cancer awareness programs in rural areas to bridge this knowledge gap.

Age at sexual debut was another key factor influencing cervical cancer knowledge. FSWs who initiated sexual activity at age 20 or older (AOR: 1.15) demonstrated higher knowledge levels compared to those with an earlier sexual debut. A possible explanation is that delaying sexual debut may provide more years of formal education and greater exposure to health-related information before becoming sexually active. In contrast, early sexual debut has been linked to lower educational attainment and reduced access to preventive health knowledge, potentially contributing to lower cervical cancer awareness in this subgroup.

Our findings align with a study in Eastern Ethiopia[26] which found that older age was associated with higher cervical cancer knowledge. Similarly, a study in Southern Ethiopia [27] reported a positive association between older age and cervical cancer awareness. Conversely, another study [28] found that individuals aged 15–19 years were less likely to have cervical cancer knowledge.

Alcohol consumption also played a role in cervical cancer knowledge. Non-drinkers had slightly higher knowledge levels compared to those who consumed alcohol (AOR: 1.08). This finding is consistent with a previous study done in China by Li and Sun, suggesting that alcohol use is associated with reduced health-seeking behaviours and limited engagement in preventive health services. [29]. Alcohol consumption may serve as a barrier to participation in health education programs, highlighting the importance of integrating harm reduction strategies into cervical cancer awareness initiatives for FSWs.

The age at which FSWs began engaging in commercial sex was significantly associated with cervical cancer knowledge. Those who entered sex work at 25 years or older had higher odds of possessing greater knowledge (AOR: 1.38). This may be attributed to increased maturity, accumulated life experiences, and previous interactions with healthcare services, which could enhance exposure to cervical cancer education. In contrast, younger entrants into commercial sex work may have had fewer opportunities to access health information, highlighting the need for targeted educational interventions for this subgroup.

To the best of our knowledge, no previous studies have specifically examined this association. Hence, this finding adds new insight into the relationship between age at entry into sex work and cervical cancer knowledge. Further research is needed to explore this association in different contexts and to better understand the underlying mechanisms that may contribute to this trend.

## Conclusion

Despite high awareness of cervical cancer among FSWs, comprehensive knowledge about its risk factors, symptoms, and prevention remains low. These findings highlight the urgent need for targeted educational interventions that consider differences in residence, age at sexual debut, age at starting commercial sex, and alcohol consumption.

## Abbreviations

AOR: Adjusted Odds Ratio
APR: Adjusted Prevalence Ratio
CI: Confidence Interval
COR: Crude Odds Ratio
FSWs: Female Sex Workers
HPV: Human Papillomavirus
KCMC: Kilimanjaro Christian Medical Centre
KCMUCo: Kilimanjaro Christian Medical University College
RDS: Respondent Driven Sampling
SD: Standard Deviation
SDGs: Sustainable Development Goals
SEM: Socioecological Model
SPSS: Statistical Package for the Social Sciences
STIs: Sexually Transmitted Infections
WHO: World Health Organization

## Declaration

### Ethical Approval

This study adhered to all ethical standards. Ethical clearance (No. PG02/2024) was obtained from Kilimanjaro Christian Medical University College (KCMUCo) and the National Health Research Ethics committee (NatHREC) (NIMR/HQ/R.8a/Vol.IX/4700). Additionally, approval letters—DC.109/228/01/K, MMC/A.40/13/VOL.V/138, and MDC/M.10/18/VOL.III/17—were secured from regional, Moshi Municipal, and Moshi District authorities, respectively.

### Patient consent for publication

All participants consented to the publication of this work

### Availability of data and materials

The dataset for this study is available upon request to the corresponding author.

### Authors Contribution

1. Gumbo D. Silas – Study conceptualization, design, data collection, analysis, report writing, and manuscript writing.
2. Federica Inturrisi – Manuscript review.
3. Innocent H. Uggh – Manuscript review.
4. Patricia Swai – Study conceptualization, and supervision.
5. Karen Yeates – Manuscript review.
6. Melinda Chelva – Data collection .and manuscript review.
7. Nicola West – Manuscript review.
8. Bariki Mchome – Study conceptualization and Manuscript review.
9. Alma R. Nzunda – Data collection and manuscript review.
10. Gaudensia Olomi – Data collection and manuscript review.
11. Prisca Marandu – Data collection and manuscript review.
12. Leah Mmari – Data collection and manuscript review..
13. Happiness Kilamwai – Data collection and manuscript review.
14. Blandina T. Mmbaga – Study conceptualization, funding acquisition, mentorship and manuscript review.
15. Alex Mremi – Study conceptualization, supportive supervision.

## Data Availability

All data produced in the present study are available upon reasonable request to the authors

## Acknowledgement

We would like to express our heartfelt gratitude to all the participants for generously sharing their time and experiences. A special thank you goes to KCRI, KCMUCo, and TAWREF for their essential support and collaboration throughout this study.

## Declaration of AI assistance

We declare that AI technology (ChatGPT) was partially employed to enhance the readability and language of this work, without altering the core ideas or content. All research tasks, data analysis, findings, and interpretations presented in this document are solely based on our original data and were conducted independently by us.

